# Higher levels of myelin are associated with higher resistance against tau pathology in Alzheimer’s disease

**DOI:** 10.1101/2022.03.02.22271746

**Authors:** Anna Rubinski, Nicolai Franzmeier, Anna Dewenter, Ying Luan, Ruben Smith, Olof Strandberg, Rik Ossenkoppele, Martin Dichgans, Oskar Hansson, Michael Ewers, Alzheimer’s Disease Neuroimaging Initiative (ADNI)

**Affiliations:** Institute for Stroke and Dementia Research, University Hospital, LMU Munich, Germany; Department of Neurology, Skåne University Hospital, Lund, Sweden; Clinical Memory Research Unit, Department of Clinical Sciences Malmö, Lund University, Lund, Sweden; Alzheimer Center Amsterdam, Department of Neurology, Amsterdam Neuroscience, Vrije Universiteit Amsterdam, Amsterdam UMC, Amsterdam, Netherlands; Munich Cluster for Systems Neurology (SyNergy), Munich, Germany; German Center for Neurodegenerative Diseases (DZNE), Munich, Germany; Memory Clinic, Skåne University Hospital, Malmö, Sweden

## Abstract

In Alzheimer’s disease (AD), pathologic tau gradually progresses from initially circumscribed predilection regions to closely connected cortical regions. The pattern of tau-deposition is of critical importance for the clinical expression of AD, but the factors that underlie region-dependent susceptibility and resistance to tau pathology remain elusive. Motivated by brain-autopsy findings suggesting late thinly myelinated regions are the first to develop tau pathology, we investigated whether the level of myelination in fiber-tracts and cortex is predictive of region-specific tau accumulation. To address this hypothesis, we combined MRI-derived template of normative myelin distribution with tau-PET imaging from two independent samples of AD-biomarker characterized participants. We found that higher myelinated cortical regions show lower tau-PET uptake in spatially corresponding areas and regions connected by highly myelinated fiber-tracts show lower rates of tau spreading. These findings were independent of amyloid-PET levels. Together, our findings suggest that higher myelination is an important resistance factor against tau pathology in AD.

## Introduction

Alzheimer’s disease (AD) is defined by the presence of beta-amyloid (Aβ) plaques and tau neurofibrillary tangles in the brain. The increase in tau pathology is closely associated with neurodegeneration and symptomatic worsening in AD ^1-3^, demonstrating a key role of tau in the development of dementia. Accumulation of tau pathology during the course of AD occurs gradually in a highly regional and connectivity-dependent manner, as fibrillar tau pathology typically starts in circumscribed brain areas and subsequently progress to anatomically connected brain areas ^4^. Entorhinal cortex is the prevailing epicenter of early fibrillar tau deposition ^5-7^, from where pathologic tau subsequently progresses to connected higher cortical areas including the medial frontal and posterior parietal cortex ^8-12^. Even at advanced disease stages of globally distributed pathologic tau, some brain regions including somatosensory and the primary motor cortex remain mostly spared ^5^. Together, these findings suggest a highly region-dependent susceptibility to tau-pathology that spreads within a network of closely connected brain regions. However, the biological substrates that underlie the region-specific vulnerability to develop tau pathology remain elusive ^13^. Their identification could provide key insights into endogenous factors of resistance against the development of fibrillar tau and may be useful for patient-tailored prediction of disease progression.

Here we investigated whether regional differences in the level of myelination are associated with lower regional fibrillar tau development. Myelin constitutes the lipid-rich insulating membrane that ensheaths the nerve fibers ^14^. Heterochronicity of nerve-fiber myelination, which can last until the 4^th^ decade of life ^15, 16^, entails differences in the level of myelination of major fiber tracts and the connecting grey matter regions: primary sensory and motor cortices are more thickly myelinated, whereas late-maturating higher cortical levels of the temporo-parietal multimodal association cortices are thinly myelinated ^16-18^. In AD, lower myelinated fiber tracts are most vulnerable ^19-21^, where the impairment of myelin may enhance the development of pathologic tau ^22^. A regional correspondence between regions of higher myelin and lower tau pathology has been noted previously based on visual inspection of brain autopsy studies ^23^. However, whether regions of higher myelination are associated with lower levels of regional tau pathology and the progressive spreading between connected brain areas has not been tested so far.

Therefore, our first major aim was to assess whether higher myelination of cortical regions is associated with lower susceptibility of fibrillar tau pathology within these regions. To this end, we combined an MRI-derived template of myelin brain mapping ^24^ with cross-sectional PET imaging of fibrillar tau, obtained from deeply phenotyped elderly participants from two independent cohorts: the Alzheimer’s Disease Neuroimaging Initiative (ADNI) ^25^ and BioFINDER-1 (https://clinicaltrials.gov/ct2/show/NCT01208675).

Our second major aim was to test whether regional levels of myelin modulated the longitudinal spreading of fibrillar tau in the brain. We have previously showed higher covariance of tau-PET accumulation between more strongly connected regions ^9, 26^, supporting the notion that tau spreads preferentially between closely connected brain regions. Here we tested the hypothesis that the level of myelination of the fiber tracts modulates the connectivity-based spreading of tau. Specifically, we hypothesized that higher myelination of fiber tracts is associated with disproportionally lower tau-PET increase in the connected brain regions. In order to address this hypothesis, we combined the myelin maps with MRI assessed structural and functional connectomes from the human connectome project (HCP) to predict longitudinal tau-PET accumulation over a time period of 1 – 4 years across both samples. Because higher levels of amyloid-PET are a major risk factor of increased cortical tau-PET, we assessed in a sensitivity analyses, whether any associations between myelin and tau-PET can be attributed to amyloid-PET levels. Our study thus investigates in a comprehensive manner in two independent samples myelin as a potentially protective factor against the progression of tau pathology.

## Results

### Study cohorts

We included a total of 612 participants consisting of non-demented and demented individuals with biomarker evidence of AD including elevated amyloid-PET accumulation (Aβ+, ADNI: n=275; BioFINDER-1: n=102) and cognitively normal (CN) controls without elevated amyloid-PET and tau-PET (Aβ-/Tau-, ADNI: n=199; BioFINDER-1: n=36, see Table 1).

**Table 1:** Sample demographics, mean (SD)

| **ADNI** |  |  |  |  |  |
| --- | --- | --- | --- | --- | --- |
|  | **CN Aβ-/Tau-**<br>**(n=199)** | **CN Aβ+**<br>**(n=119)** | **MCI Aβ+**<br>**(n=97)** | **AD dementia**<br>**(n=59)** | ***p-value*** |
| Age, years | 70.88 (6.40) | 74.64 (7.39) * | 74.77 (7.40) * | 76.20 (9.33) * | <0.001 |
| Sex, M/F | 72 / 127 | 44 / 75 | 51 / 46 | 31 / 28 | 0.011 |
| Education, years | 16.78 (2.28) | 16.68 (2.38) | 16.11 (2.46) | 15.44 (2.41) *† | <0.001 |
| MMSE | 29.19 (1.07) | 28.91 (1.36) | 27.49 (2.19) *† | 22.19 (3.96) *†‡ | <0.001 |
| APOE ε4 carriers -/+<sup>1</sup> | 136- / 51+ | 54- / 60+* | 29- / 53+* | 18- / 32+* | <0.001 |
| Mean tau-PET follow-up, years<sup>2</sup> | 1.72 (0.53) | 1.64 (0.57) | 1.45 (0.71) | 1.37 (0.49) | 0.299 |
| **BioFINDER-1** |  |  |  |  |  |
|  | **CN Aβ-/Tau-**<br>**(n=36)** | **CN Aβ+**<br>**(n=30)** | **MCI Aβ+**<br>**(n=26)** | **AD dementia**<br>**(n=46)** | ***p-value*** |
| Age, years | 73.86 (7.27) | 73.80 (7.44) | 71.73 (9.72) | 71.43 (7.34) | 0.274 |
| Sex (M/F) | 19 / 17 | 12 / 18 | 18 / 8 | 25 / 21 | 0.187 |
| Education, years | 12.42 (3.80) | 11.93 (3.72) | 12.21 (3.93) | 12.22 (3.75) | 0.928 |
| MMSE | 28.86 (1.10) | 28.83 (1.15) | 25.58 (3.06) *† | 20.96 (4.87) *†‡ | < 0.001 |
| APOE ε4 carriers -/+ | 29- / 7+ | 8- / 22+* | 6- / 20+* | 19- / 27+* | < 0.001 |
| Mean tau-PET follow-up, years<sup>3</sup> | 2.03 (0.47) | 1.94 (0.32) | 1.82 (0.12) | 1.87 (0.34) | 0.470 |

Abbreviations: Aβ, amyloid-beta; AD, Alzheimer's disease; APOE, Apolipoprotein E; CN, cognitively normal; F, female; M, male; MCI, mild cognitive impairment; MMSE, Mini-Mental State Exam.
1 = available for 187 CN Aβ-/Tau-, 114 CN Aβ+, 82 MCI Aβ+, and 50 AD dementia.
2 = subsample of 35 CN Aβ-/Tau-, 60 CN Aβ+, 39 MCI Aβ+, and 24 AD dementia.
3 = subsample of 16 CN Aβ-/Tau-, 14 CN Aβ+, 7 MCI Aβ+, and 18 AD dementia.
\*Significantly different from CN Aβ-/Tau-, † significantly different from CN Aβ+, ‡significantly different from MCI Aβ+, §significantly different from AD dementia. (Significant after applying a Bonferroni-corrected α-threshold of 0.0125)

### Higher cortical myelination is associated with lower tau-PET

Our first aim was to test in a cross-sectional analysis whether higher myelin levels in grey matter regions were associated with lower tau-PET uptake in spatially corresponding regions. To this end, we first applied a 200 region-of-interest (ROI) cortical brain-parcellation atlas (Figure 1A) to a spatially normalized MRI-based myelin-water-fraction (MWF) template from cognitively normal healthy adults (n=50, mean age 25 years) ^24^ in order to extract normative MWF ROI values from cortical brain regions (Figure 1B). Next, we obtained subject-level tau-PET SUVRs from the same 200 ROI based on the tau-PET scans in the ADNI and BioFINDER-1 cohorts (Figure 1C). The tau-PET ROI SUVRs were weighted by tau-positivity probability ^7, 12^ in order to reduce the influence of off-target background binding (henceforth called tau-PET scores, see supplementary Figure 1). We used spatial correlation to assess whether those grey matter ROIs with higher MWF levels show lower tau-PET scores in AD (Figure 1D). As hypothesized, we found that higher ROI-MWF was associated with lower tau-PET scores in corresponding grey matter ROIs for Aβ+ participants in both ADNI (rho=-0.267, p<0.001; Figure 2A) and BioFINDER-1 (rho=-0.175, p=0.013; Figure 2B). The analysis in the CN Aβ-/Tau-participants was not significant both in ADNI (rho=-0.13, p=0.067; Figure 2A) and BioFINDER-1 (rho=0.027, p=0.7; Figure 2B), suggesting that the association between MWF and tau-PET scores is robust only in the group with biomarker evidence of AD.

**Figure 1:**
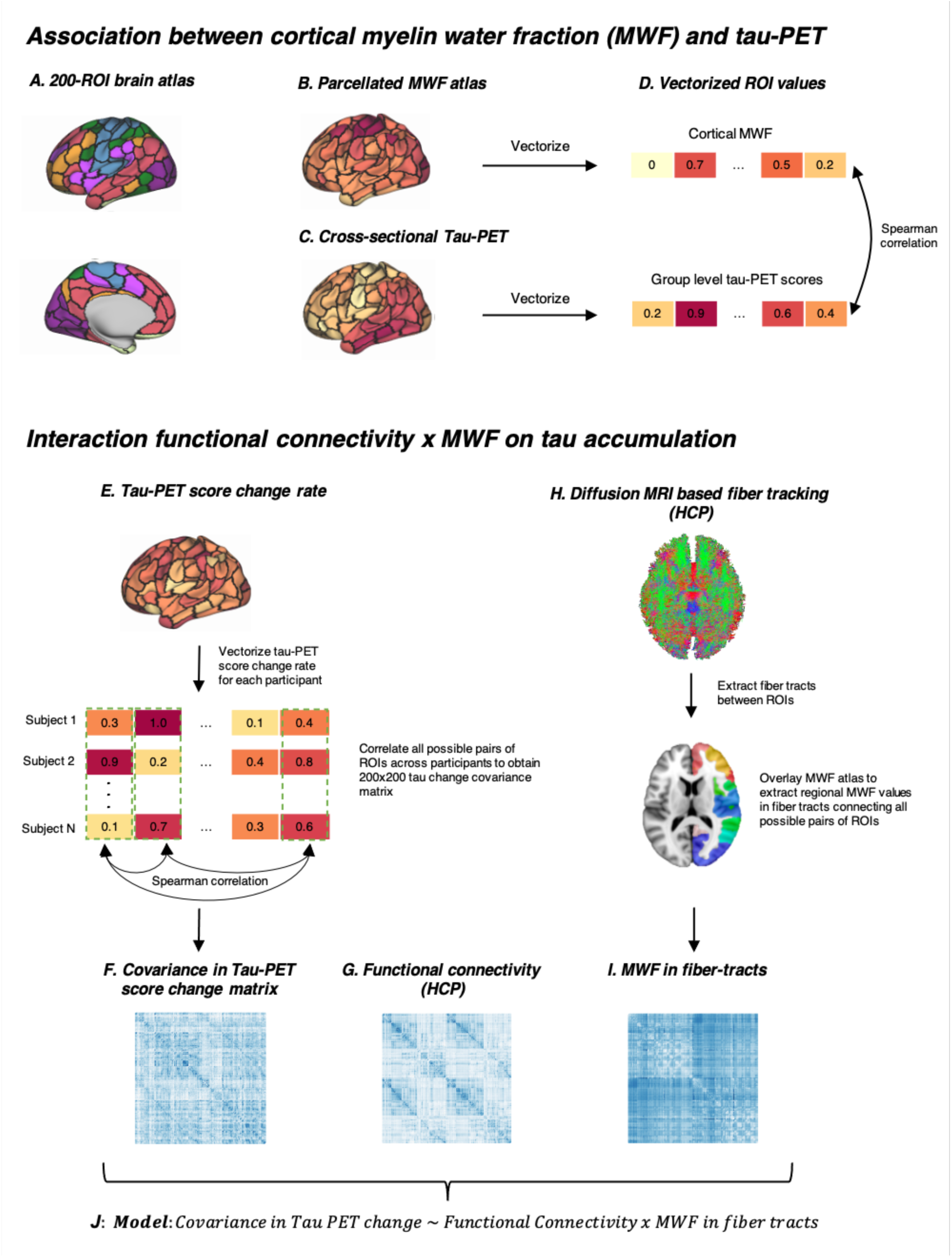
Analysis flow chart. (A) Surface rendering of the 200-ROI brain atlas, based on which we estimated (B) cortical WMF and (C) group-level tau-PET scores, which were further vectorized (D) and spatially correlated. (E) The same brain atlas shown in A was applied to each participants tau-PET score change rates, individual values were vectorized and all possible pairs of ROIs across participants were correlated to obtain a covariance in tau-PET score change matrix (F). (G) Using the 200 ROI brain atlas shown in A, resting-state fMRI functional connectivity was assessed on 100 participants of the human connectome project (HCP). (F) Diffusion MRI from the HCP was used to estimate the fiber-tracts between each pair of ROIs in the brain atlas shown in A. (I) The MWF atlas was overlayed to extract regional MWF values of underlying fiber-tracts. (J) Linear regression analysis was performed with the covariance in tau-PET score change as the dependent variable, and the interaction of functional connectivity by MWF in fiber-tract as the predictor. In a sensitivity analyses we further controlled the above-mentioned analyses for regional amyloid-PET levels or the covariance in amyloid-PET change (not shown).

**Figure 2:**
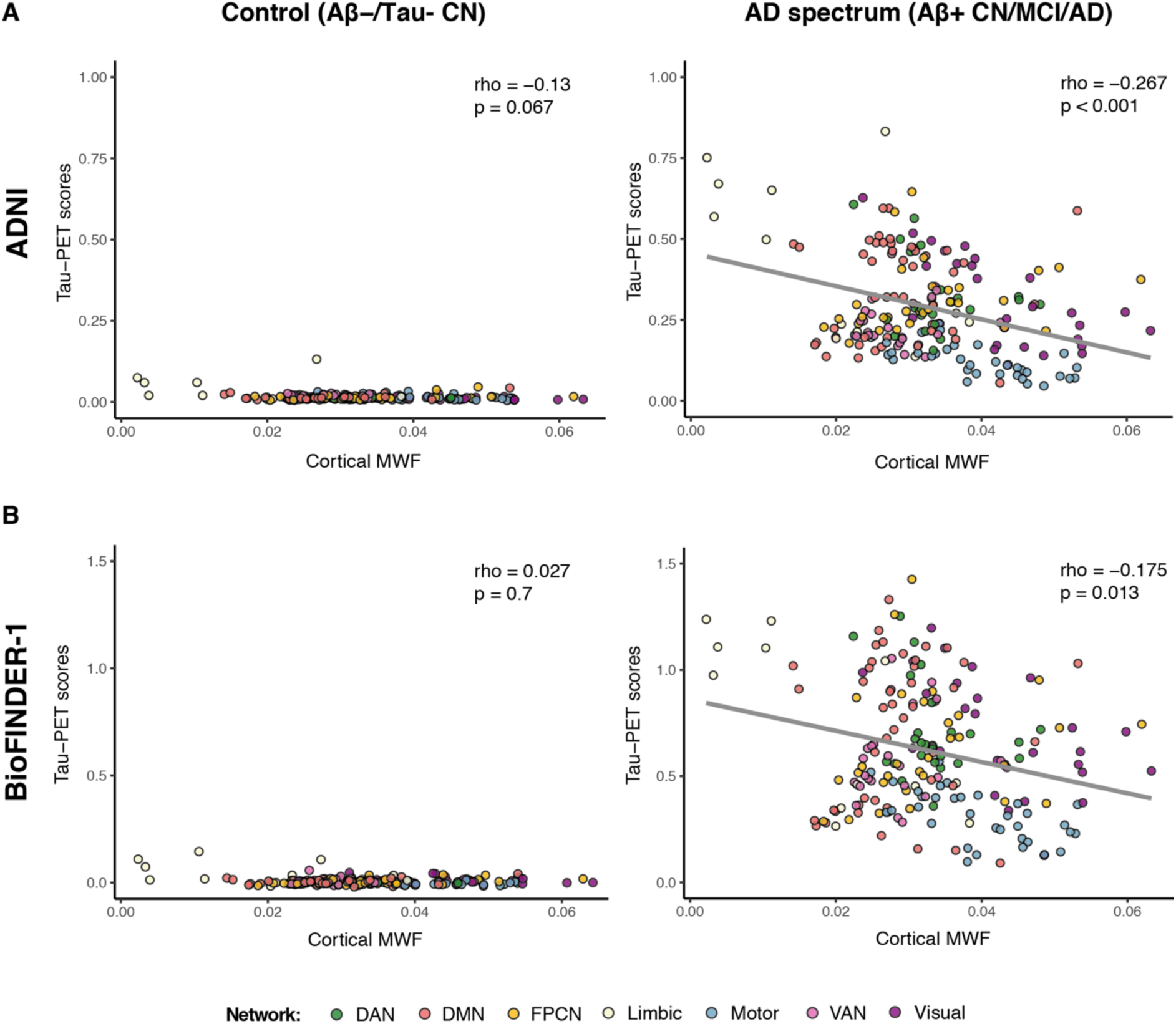
Association between cortical MWF and baseline tau-PET scores. Scatterplots showing the association between ROI levels of MWF and tau-PET scores for controls (CN Aβ-/Tau-; left column) and AD spectrum (Aβ+ participants; right column) from the ADNI (A) and BioFINDER-1 (B) cohorts. The coloring indicates for each ROI the major functional network it belongs to. DAN = Dorsal Attention Network; DMN = Default-Mode Network; PFCN = Fronto-Parietal Control Network; VAN = Ventral Attention Network; MWF = Myelin Water Fraction.

As a sensitivity analysis we repeated the analysis using tau-PET SUVRs instead of tau-PET scores. We found that higher ROI-MWF was associated with lower tau-PET SUVRs in corresponding grey matter ROIs for Aβ+ participants in both ADNI (rho=-0.348, p<0.001; Supplementary Figure 2A) and BioFINDER-1 (rho=-0.255, p<0.001; Supplementary Figure 2B).

### Higher myelination of fiber tracts attenuates rates of longitudinal tau-PET increase in connected regions

We and others previously reported that tau-PET preferentially progresses between closely connected brain regions ^9^. Here, we tested whether myelination of fiber tracts modulates the rate of connectivity-dependent tau-PET accumulation in Aβ+ participants. We addressed this hypothesis in a subset of Aβ+ participants who had longitudinal tau-PET (ADNI: n=123, mean FU interval = 1.53 [0.69-3.95] years; BioFINDER-1: n=39, mean FU interval = 1.87 [1.21-2.78]). Adopting our previously established approach to assess connectivity-based tau accumulation ^9, 26^, we first estimated the functional connectivity between each of the ROIs (Figure 1G). Next, in order to determine whether functionally connected regions show similar rates of tau accumulation, we computed the covariance in tau-PET score change (i.e, the level of similarity in tau change between two regions; Figure 1E-F). We then determined the level of myelin in the fiber-tracts connecting each pair of ROIs to test whether myelin levels modulate the association between the functional connectivity and the change in tau-PET in the connected ROIs. To this end, we obtained the normative fiber-tract myelin levels from the same MWF template as used before, this time masked by an ROI-to-ROI structural connectivity matrix based on diffusion MRI data from 100 healthy individuals assessed in the HCP (Figure 1H-I). In a linear regression analysis, we tested the interaction functional connectivity by MWF in fiber-tracts as predictors of the covariance in tau change matrix (Figure 1J). We found a significant interaction of functional connectivity by fiber-tract MWF on covariance in tau change in both ADNI (β=-0.185, p<0.001; Figure 3A) and BioFINDER-1 (β=-0.166, p<0.001; Figure 3B), where regions that were connected by higher myelinated fiber tracts showed a lower association between functional connectivity and the rate of change in tau-PET scores. These associations were not significant in the control groups (ADNI: β=0.027, p=0.39; BioFINDER-1: β=-0.026, p=0.43; Figure 3A-B). These results suggest that in the groups with biomarker evidence of AD, myelin is associated with an attenuated rate of connectivity-dependent tau-PET increase over time.

**Figure 3:**
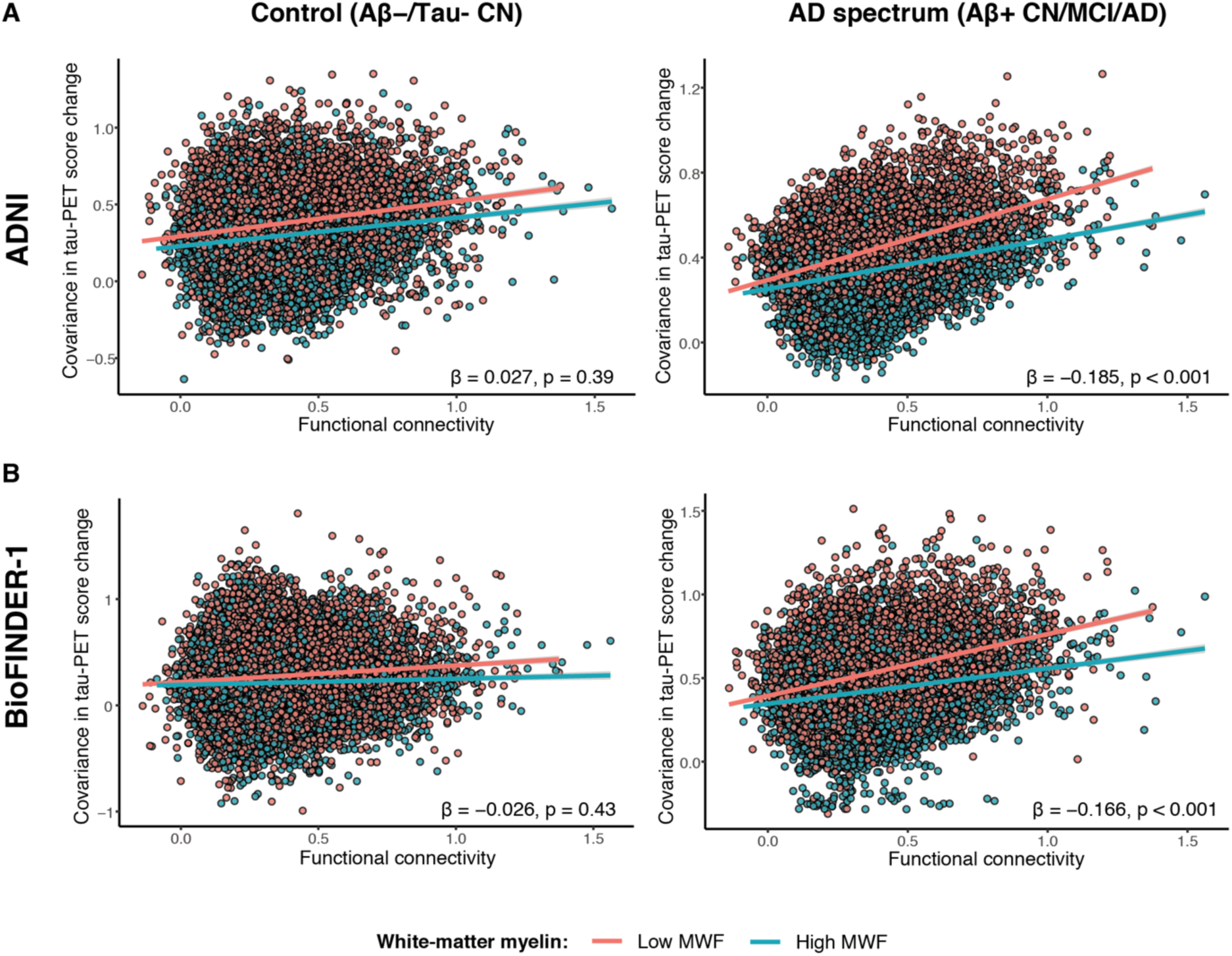
Interaction between functional connectivity and MWF in fiber-tracts on covariance in tau-PET score change. Regression plots illustrating covariance in tau-PET change as a function of both functional connectivity and MWF in fiber-tracts (binarized by median spit) in the controls (CN Aβ-/Tau-; left column) and AD spectrum (Aβ+ participants; right column) groups of the ADNI (C) and BioFINDER-1 (D) cohorts. Red line is the regression line for participants with values < median MWF, and the blue regression line is for participants with values > median MWF. For the statistical analyses MWF was used as a continues measure and was stratified to high and low only for illustrational purposes. MWF = Myelin Water Fraction.

### The associations between tau and MWF are not driven by amyloid

Lastly, in a sensitivity analyses we tested whether the above-mentioned associations were driven by regional amyloid pathology. We addressed this in the larger ADNI sample in whom amyloid-PET was acquired in close temporal proximity to baseline tau-PET (mean interval = 20.5 [1-156] days) in 168 Aβ+ participants.

First, we tested the association between cortical MWF and tau-PET scores, controlling for regional amyloid-PET SUVRs. We found that higher cortical MWF was associated with lower tau-PET scores in corresponding grey matter ROIs independent of amyloid-PET (rho=-0.21, p=0.003).

Next, we tested whether the association between higher fiber-tract MWF and lower connectivity-dependent longitudinal tau-PET accumulation remained significant. When controlling for the covariance in amyloid-PET change, the interaction functional connectivity by MWF in fiber tracts on covariance in tau change remained significant (β=-0.07, p=0.005), suggesting that the association between levels of higher myelin and lower tau-PET accumulation was not dependent on differences in Aβ.

## Discussion

Our first major finding in two independent cohorts of AD-biomarker positive samples showed that higher levels of myelin as assessed by MWF were consistently associated with lower tau-PET uptake in corresponding grey matter regions. This result suggests that brain regions with higher myelin levels are less prone to accumulate fibrillar tau. Our second major finding concerned the longitudinal progression of tau-PET between connected brain areas. We revealed that higher MWF levels in fiber-tracts were associated with lower connectivity-dependent rates of tau-PET increases. The associations remained robust when controlling for amyloid-PET accumulation, suggesting that the associations between myelin and tau-PET were not merely a function of differences in Aβ. This result suggests that higher fiber-tract myelination is associated with attenuated spreading of fibrillar tau in the brain. Although we caution against a causative interpretation, our current findings suggest that higher regional levels of myelin are associated with higher resistance against the susceptibility and rate of progression of fibrillar tau.

We combined for the first-time high-resolution MRI-assessed mapping of myelin, connectomics, and subject-level tau-PET imaging in two independent samples of deeply characterized elderly individuals with biomarker evidence of AD. This allowed us to reveal the close spatial association between higher myelin levels and lower cross-sectional and longitudinal increase in tau-PET accumulation. Our findings significantly advance previous qualitative brain autopsy studies that suggested an association between myelin and tau pathology ^21, 23, 27^, but were limited to qualitative visual inspection of sparsely sampled post-mortem data of tau pathology and myelin maps ^23^. Here, using a longitudinal tau-PET approach, we show that higher fiber-tract myelination is associated with lower connectivity-dependent rate of tau-PET progression. We thus advance previous findings on the association between connectivity-based prediction of faster tau-PET accumulation in regions which are closely connected ^9, 12, 28, 29^, demonstrating that such connectivity-related increases in tau-PET accumulation are attenuated for regions connected by higher myelinated fiber-tracts. Furthermore, we show that the associations between myelin and tau-PET accumulation remained significant after accounting for regional amyloid-PET, suggesting that our main results on tau-PET were not modulated by amyloid deposition. We note that these results do not rule out the possibility that regional premorbid differences are related to differences in amyloid plaque deposition, rather these results demonstrate that any association between myelin and Aβ deposition cannot account for the association with tau-PET. Together, our findings strongly support the notion that higher myelination is associated with lower susceptibility to tau accumulation and contribute to explain why some regions are highly vulnerable to tau pathology whereas other are relatively spared.

The mechanisms that may underlie a protective effect of higher myelin on lower tau pathology are poorly understood. A possible explanation is that lower myelinated axons are more prone to myelin damage, where subsequent remyelination processes entails higher phosphorylation of tau and subsequent formation of fibrillar tau ^30, 31^. Experimentally induced myelin damage leads to the activation of tau-targeting kinases entailing hyperphosphorylation of tau ^22, 32^, and could trigger further spreading of tau pathology ^33, 34^. At the molecular level, the Fyn kinase, which is a key signaling molecule that binds tau in axonal microtubules ^35^, becomes activated during remyelination efforts triggered by myelin disturbances ^36^. In mouse models of tauopathies, blocking Fyn kinase completely prevented the formation of neurofibrillary tangles ^37^. Together, these results suggest the possibility that myelin-triggered Fyn kinase may play a role in the formation of tau tangles in AD. Myelin damage has been observed previously in histochemical and neuroimaging studies in AD ^38-41^, and recent single cell studies provide converging results showing that alterations in oligodendrocytes, a cell-type involved in myelination, are prominent in AD ^42, 43^. Recent studies in transgenic mouse models of amyloid pathology suggest that acute demyelination may occur *upstream* of amyloid deposition by interfering with a TREM2-related microglia activation signature, thus leading to enhanced amyloid deposition ^44^. Thus, more vulnerable, thinner myelinated regions may engage in dysfunctional microglial activity resulting in less efficient removal of AD pathology. Whether such a myelin-related interference with microglial activation upstream of Aβ extends to the development of fibrillar tau remains to be tested. In summary, regions with higher myelination may be less vulnerable to late-life myelin alterations and thus a pathological cascade that includes the development of AD pathologies.

Our findings have important clinical implications. First, our results suggest that myelin adds to the connectivity-based prediction of regional increases in tau-PET ^9, 12^, and may thus contribute to patient-tailored outcome parameters on treatment effects on tau-PET changes ^45^. Second, the current findings suggest myelin to be a potential target for the prevention and treatment of AD. Myelination is a druggable target ^46, 47^, and thus pharmacological stimulation of myelination is a putative therapeutic target in AD. Clemastine, a licensed H1 histamine, has been recently shown to enhance the differentiation of oligodendrocyte precursor cells (OPC) and myelination ^48, 49^, where clinical trials repurposing clemastine for the treatment of multiple sclerosis have shown improvement of symptoms ^50^. Treatment of transgenic mouse model of Aβ with clemastine showed alleviated OPC and myelin loss, reduced Aβ deposition and ameliorated memory loss ^31, 51^. Together, these results suggest that myelin is a putative drug target in AD and encourage future studies to test the effect of myelin treatment on tau pathology.

### Limitations

Several caveats should be considered when interpreting our results. First, current MRI acquisition sequences are imperfect measures of myelin and are susceptible to iron content in the brain tissue ^52^. However, we employed an MR-based myelin water imaging ^24^, which has been extensively validated by histopathology ^53, 54^ and shows one of the strongest associations with post-mortem assessed myelin levels compared to alternative MRI measures of myelin ^55^. Also, we avoided a potential confounding of the MRI myelin signal with disease-related iron and white matter alterations by using a myelin water imaging template that was obtained in healthy individuals ^24^. Therefore, the current findings are unlikely to be driven by spurious associations with off-target MRI signals. Second, we did not assess myelin changes in the AD patients. However, demyelination has been observed in association with core AD pathologies including amyloid plaques ^40^ and fibrillar tau ^38^. Hence, myelin changes in AD may have altered the rate of change in tau-PET. Patient-specific levels of myelin measurement was not available in the current study, but the hypothesis is that regions with lower levels of myelin are more prone to undergo demyelination which then triggers the development of tau pathology.

In summary, our findings show for the first time that higher myelination is associated with slower tau progression in AD. Our results demonstrate that myelin maps inform connectome-based prediction of pathology-spreading models, and thus also pave the way for future clinical studies in which the individual interregional differences in myelination can be assessed for patient-tailored prediction of disease progression.

## Methods

### Sample

The current study included participants from two large cohort studies: the Alzheimer’s disease neuroimaging initiative (ADNI) and the Swedish BioFINDER-1 study. For our main analyses, participants were selected based on the availability of baseline T1-weigthed MRI and [^18^F]-AV1451 (flortaucipir) tau-PET. For a sensitivity analyses, for ADNI participants we also acquired all available [^18^F]-AV45 (Florbetapir) amyloid-PET data.

For ADNI, Aβ status (Aβ-/+) was determined based on global amyloid-PET levels, where abnormally elevated amyloid deposition (Aβ+) was defined as a cerebellum-normalized global standardized uptake value ratio (SUVR) cut-off > 1.11 for [^18^F]AV45-Florbetapir-PET and global SUVR > 1.08 for [^18^F]Florbetaben-PET as established previously ^56^. Participants were clinically diagnosed as cognitively normal (CN, MMSE > 24, Clinical Dementia Rating (CDR) = 0, nondepressed), mildly cognitively impaired (MCI, MMSE > 24, CDR = 0.5, objective memory loss on the education adjusted Wechsler Memory Scale II, preserved activities of daily living), or demented (AD, MMSE of 20 to 26, CDR > 0.5, NINCDS/ADRDA criteria for probable AD). We included a group of participants within the AD spectrum consisting of 119 CN Aβ+, 97 MCI Aβ+, and 59 Aβ+ AD dementia participants. As a control sample we included 199 CN Aβ-/Tau-individuals. Tau negativity was established based on a temporal meta-ROI (including the amygdala, entorhinal cortex, fusiform, parahippocampal, and inferior temporal and middle temporal gyri) cut-off < 1.29, which was shown to discriminate non-AD from AD participants ^57^. Ethical approval was obtained by the ADNI investigators at each participating ADNI site, all participants provided written informed consent.

For BioFINDER-1, Aβ status was determined based on global [^18^F]Flutemetamol-PET levels, where abnormally elevated amyloid deposition was defined as a global SUVR cut-off > 0.575 as described previously ^58^. The inclusion and exclusion criteria and diagnostic criteria for BioFINDER-1 have been published previously ^59^. We included a control sample of 36 CN Aβ-/Tau-, and a group of participants within the AD spectrum consisting of 30 CN Aβ+, 26 MCI Aβ+, and 46 Aβ+ AD dementia participants. All BioFINDER-1 participants gave written informed consent to participate in the study prior to inclusion in the study. Ethical approval was given by the ethics committee of Lund University, Sweden. Imaging procedures were approved by the Swedish Medical Product Agency and the Radiation Safety Committee at Skåne University Hospital, Sweden.

### MRI and PET acquisition and preprocessing in ADNI

In ADNI, structural MRI data was acquired on 3T scanning platforms using T1-weighted MPRAGE sequences using unified scanning protocols across sites (image acquisition procedures can be found on: http://adni.loni.usc.edu/methods/mri-tool/mri-analysis/).

Tau-PET was assessed with a standardized protocol using 6 × 5 min frames, 75-105 min post-injection of [^18^F]-AV1451. Similarly, amyloid-PET was acquired in 4 × 5 min frames, 50-70 min post injection of [^18^F]-AV45. The dynamically acquired frames were coregistered and averaged, and further standardized with respect to the orientation, voxel size and intensity by the ADNI PET core to produce uniform single tau-PET ^60^.

T1-weigthed MRI images and PET data were preprocessed using the Advanced Normalization Tools (ANTs) toolbox (http://stnava.github.io/ANTs/). First, PET images were rigidly co-registered to the participant’s T1-weigthed MRI image in native space. Using the ANTs cortical thickness pipeline, T1-weigthed images were bias field corrected, brain extracted, and segmented into grey matter, white matter, and cerebrospinal fluid tissue maps. Using ANTs high-dimensional warping algorithm ^61^ the preprocessed T1-weigthed MRI images were further non-linearly normalized to MNI space. By combining the normalization parameters, we further transformed the Schaefer 200 ROI cortical brain atlas parcellation ^62^ and the reference region for intensity normalization of PET images from MNI space to native space. Subsequently, the Schaefer parcellation and the reference region were masked with subject-specific grey matter masks that were binarized at a probability threshold of 0.3.

PET SUVR images were computed by intensity normalizing PET images to the mean tracer uptake of the inferior cerebellar grey matter for tau-PET data, or to the mean tracer uptake of the whole cerebellum for amyloid-PET data, following previous recommendations ^63, 64^. Mean PET SUVR values were extracted for each subject for the 200 cortical ROIs (Figure 1C).

### MRI and PET acquisition and preprocessing in BioFINDER-1

In BioFINDER-1, T1-weigthed MPRAGE (1mm isotropic, TR/TE=1900/2.64ms) and FLAIR images (0.7 × 0.7 × 5 mm^3^, 23 slices, TR/TE=9000/81ms) were acquired for all participants on a 3T Siemens Skyra scanner (Siemens Healthineers, Erlangen, Germany).

Tau-PET was acquired 80-100 minutes after bolus injection of [^18^F]AV1451 on a GE Discovery 690 PET scanner (General Electric Medical Systems, Milwaukee, WI, USA).

The image data was processed by the BioFINDER-1 imaging core using a previously described pipeline developed at Lund University ^65^. In brief, MRI images were skull stripped using the combined MPRAGE and FLAIR data, segmented into grey and white matter and non-linearly normalized to MNI space. PET images were attenuation corrected, motion corrected, summed, and coregistered to the MRI images.

In line with the ADNI data, SUVR images were computed by using the inferior cerebellar grey matter as a reference region, and mean tau-PET SUVR values were extracted for each subject for the 200 cortical ROIs.

### Transforming tau-PET SUVRs to tau-PET scores

The AV1451 tau-PET tracer shows off-target binding which can confound the measurement of fibrillar tau. In order to enhance the specificity of the tau-PET tracer to assess fibrillar tau, we employed gaussian mixture modeling to tau-PET data to separate target signal from background signal as previously described ^7, 12^. The underlying rationale is that the variability of regional tau-PET values arises from two sources including off-label bindings observed mostly in regions free of fibrillar tau and those with abnormally increased fibrillar tau (such as present in Aβ+ participants), resulting in a bimodal distribution of ROI values centered around a mean of off-label tracer binding and a distribution of ROI values centered around a mean of on-target tracer binding. For a given participant’s ROI value, a tau-positivity probability score (i.e. the probability that the tau-PET ROI value reflects abnormal fibrillar tau) can be calculated based on the percentiles of the off-label and target ROI value distributions. In the current study we applied gaussian mixture modelling to 200-ROI tau-PET SUVRs across participants separately in each study including ADNI and BioFINDER-1. We computed for each participant and ROI the probability to belong either to the off-target or on-target distribution as described previously by us ^12^. The resulting tau-positivity probabilities were then multiplied by the original tau-PET SUVRs in order to obtain tau-positivity weighted tau-PET scores.

### Assessment of covariance in tau change

Using longitudinal tau-PET data, we then computed the annual tau change within each of the 200 ROIs for each participant. This was done by computing for each ROI the baseline vs follow-up differences in tau-PET scores divided by the time (in years) between the tau scans. Covariance in tau change (the correlation between tau change levels between pair of regions) was computed by correlating regional tau-PET score change levels between each pair of ROIs across participants using Spearman’s correlation. Resulting in a single 200×200 sized covariance in tau change matrix for both cohorts. We set autocorrelations to zero and further Fisher-z transformed all correlations. Similarly, for a sensitivity analysis we computed the covariance in amyloid-PET change.

### Assessment of cortical and fiber tract MWF

Cortical and fiber tracts myelination was determined based on a normative MWF atlas derived from myelin water imaging of healthy individuals (n=50, mean age 25 years) ^24^. To determine the regional cortical distribution, we applied a neocortical 200 ROI brain parcellation Figure 1A; ^62^ to the myelin water atlas, and extracted the regional mean MWF within each of the 200 ROIs (Figure 1B). To determine the myelin content within the fiber-tracts connecting each pair of ROIs, first we obtained minimally preprocessed diffusion-weighted images (DWI) ^66^ from 100 participants of the human connectome project (HCP). We further used a multi-shell multi-tissue constrained spherical deconvolution and probabilistic tractography pipeline, as previously described ^12^. Using the same 200 ROI parcellation, we defined nodes and assigned for each pair of ROIs the reconstructed fiber tract streamlines (Figure 1H). The myelin water atlas was then applied to the reconstructed streamlines, and the mean myelin value along the streamlines between each of two regions was extracted. This resulted in a 200 × 200 matrix of the mean MWF in fiber-tracts between each pair of ROIs. From the resulting 100 MWF matrices, we computed the group-average MWF in fiber-tracts matrix (See Figure 1I).

### Assessment of functional connectivity

To determine a functional connectivity template, we downloaded spatially normalized, minimally preprocessed resting-state fMRI images from the same 100 participants of the HCP. We then applied detrending, band-pass filtering (0.01-0.08 Hz), despiking and motion correction to the HCP resting-state data. For each individual, we assessed functional connectivity as Fisher-z transformed Pearson-moment correlations between all possible ROI pairs in the Schaefer parcellation. From the resulting 100 functional connectivity matrices, we computed the group-average functional connectivity matrix (Figure 1G).

### Statistics

Within each cohort, subject characteristics were compared between groups using Kruskal-Wallis for continues measures (post hoc Dunn’s tests adjusted for multiple comparisons) or Chi-squared (χ2) tests for categorical measures.

In order to test the association between cortical MWF and group-averaged tau-PET scores in corresponding grey matter ROIs, we computed a spatial correlation using Spearman’s rank correlation (Figure 1B-D) separately for ADNI and BioFINDER-1. As sensitivity analyses, we repeated this analysis using tau-PET SUVRs as the dependent variable.

Next, we assessed whether myelination of fiber-tracts is modulating the connectivity-based spreading of tau. To that end, using linear regression we tested the interaction functional connectivity by MWF in fiber-tracts on the covariance in tau-PET score change separately for ADNI and BioFINDER-1.

Finally, in order to assess whether the observed associations in the current study are driven by regional amyloid pathology, we repeated the above-mentioned analyses controlling for baseline amyloid-PET levels or covariance in amyloid-PET change based on the analysis.

All statistical analyses were performed using R statistical software. Brain surface renderings were created in connectome workbench. All effects were considered significant when meeting an α-threshold of 0.05.

## Data Availability

The data used in this study were obtained from the Alzheimer's disease Neuroimaging Initiative and are available from the ADNI database (adni.loni.usc.edu) upon registration and compliance with the data usage agreement. Data from the BioFINDER-1 cohort are available from the authors upon request. Resting-state and diffusion-weighted data of the HCP cohort are freely available online (https://db.humanconnectome.org). MWF template is freely available online (https://sourceforge.net/projects/myelin-water-atlas/).

## Author contributions

A.R.: study concept and design, data processing, statistical analysis, interpretation of the results, writing the manuscript. N.F.: data processing, critical revision of the manuscript; A.D.: data processing, critical revision of the manuscript. Y.L.: data processing, critical revision of the manuscript. R.S.: critical revision of the manuscript. O.S.: critical revision of the manuscript. R.O.: critical revision of the manuscript. M.D.: critical revision of the manuscript. O.H.: critical revision of the manuscript. M.E.: study concept and design, interpretation of the results, writing the manuscript.

## Acknowledgements

Parts of the data used in preparation of this manuscript were obtained from the ADNI database (adni.loni.usc.edu). As such, the investigators within the ADNI study contributed to the design and implementation of ADNI and/or provided data but did not participate in analysis or writing of this paper. A complete listing of ADNI investigators can be found at the end of the manuscript.

The study was supported by the German Center for Neurodegenerative Diseases (DZNE), and the Deutsche Forschungsgemeinschaft (DFG, German Research Foundation) grant for major research instrumentation (DFG, INST 409/193-1 FUGG).

ADNI data collection and sharing for this project was funded by the ADNI (National Institutes of Health Grant U01 AG024904) and DOD ADNI (Department of Defense award number W81XWH-12-2-0012). ADNI is funded by the National Institute on Aging, the National Institute of Biomedical Imaging, and Bioengineering, and through contributions from the following: AbbVie, Alzheimer’s Association; Alzheimer’s Drug Discovery Foundation; Araclon Biotech; BioClinica, Inc.; Biogen; Bristol-Myers Squibb Company; CereSpir, Inc.; Cogstate; Eisai Inc.; Elan Pharmaceuticals, Inc.; Eli Lilly and Company; EuroImmun; F. Hoffmann-La Roche Ltd and its affiliated company Genentech, Inc.; Fujirebio; GE Healthcare; IXICO Ltd.; Janssen Alzheimer Immunotherapy Research & Development, LLC.; Johnson & Johnson Pharmaceutical Research & Development LLC.; Lumosity; Lundbeck; Merck & Co., Inc.; Meso Scale Diagnostics, LLC.; NeuroRx Research; Neurotrack Technologies; Novartis Pharmaceuticals Corporation; Pfizer Inc.; Piramal Imaging; Servier; Takeda Pharmaceutical Company; and Transition Therapeutics. The Canadian Institutes of Health Research is providing funds to support ADNI clinical sites in Canada. Private sector contributions are facilitated by the Foundation for the National Institutes of Health (www.fnih.org).

