## Supplementary figures for "Higher levels of myelin are associated with higher resistance against tau pathology in Alzheimer’s disease"

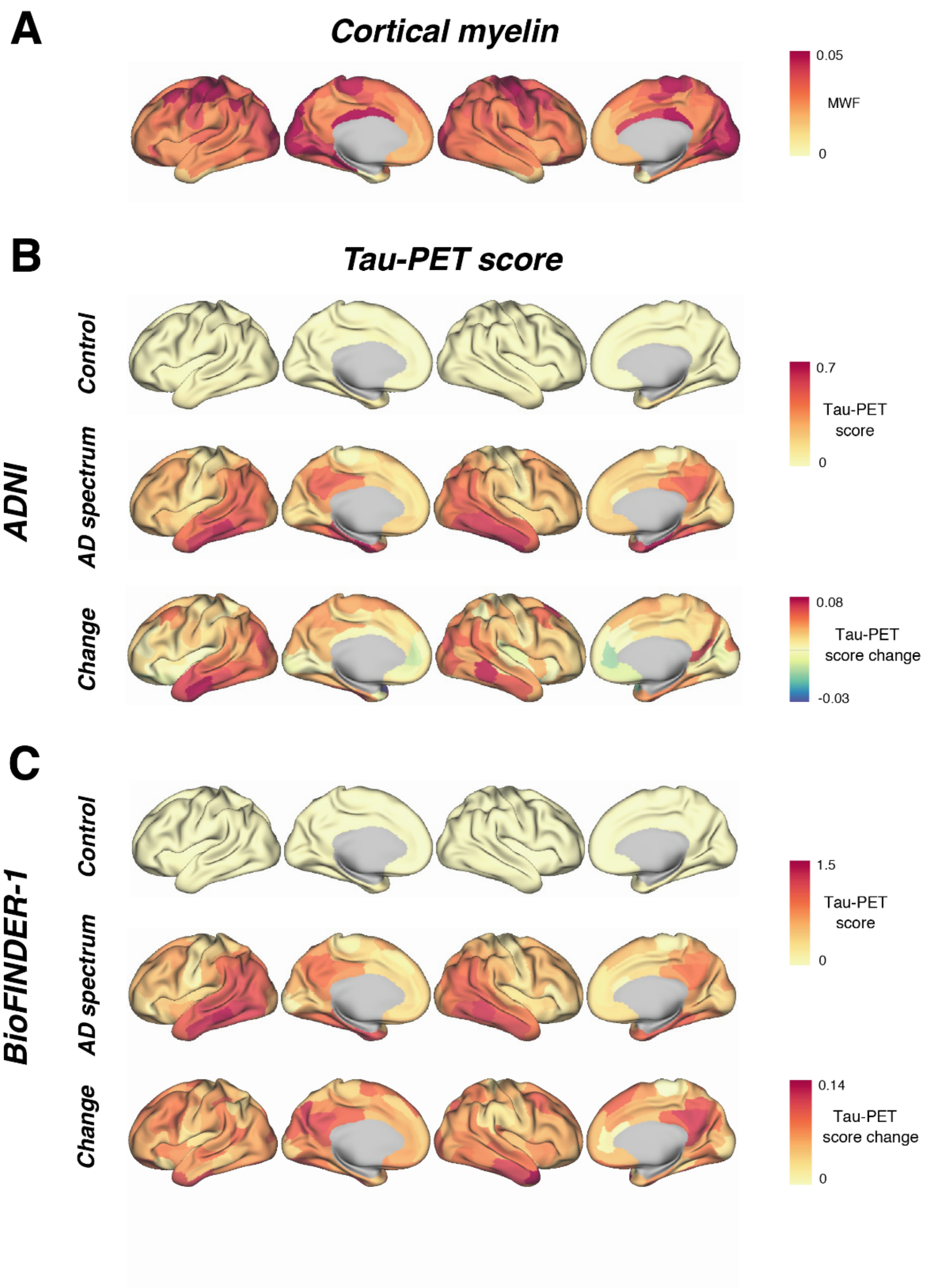


**Supplementary figure 1: Brain renderings of cortical myelin within the MWF template and tau-PET scores among controls and AD participants.**

Surface renderings of cortical myelin distribution derived from the MWF atlas of healthy individuals (A). Tau-PET scores averaged within controls (i.e. CN Aβ-/Tau-) and AD spectrum (Aβ+ participants) and change in tau-PET scores averaged within the AD spectrum for ADNI (B) and BioFINDER-1 (C) cohorts.

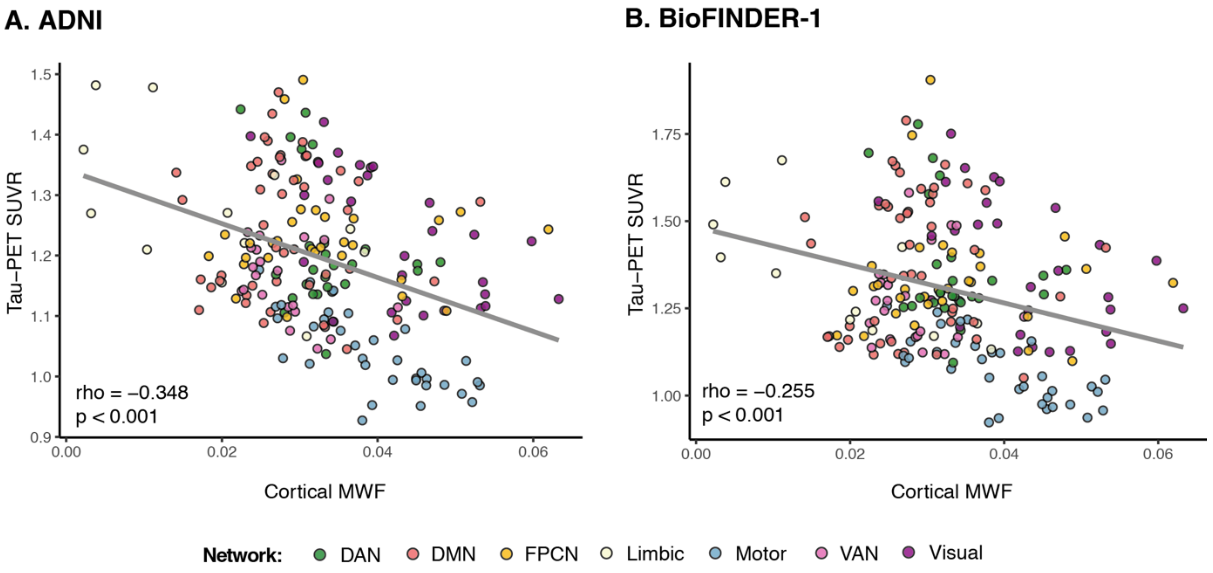


**Supplementary figure 2: Association between cortical MWF and baseline tau-PET SUVRs**
Scatterplots showing the association between ROI levels of MWF and tau-PET SUVR for the AD spectrum (Aβ+ participants) from the ADNI (A) and BioFINDER-1 (B) cohorts. The coloring indicates for each ROI the major functional network it belongs to. DAN = Dorsal Attention Network; DMN = Default-Mode Network; PFCN = Fronto-Parietal Control Network; VAN = Ventral Attention Network; MWF = Myelin Water Fraction.
